# Social Behavioral Impairments in *SYNGAP1*-Related Intellectual Disability

**DOI:** 10.1101/2023.03.11.23287144

**Authors:** Hajera Naveed, Maria McCormack, J. Lloyd Holder

## Abstract

Synaptopathies are neurodevelopmental disorders caused by genetic mutations disrupting the development and function of neuronal synapses. We administered the validated Social Responsiveness Scale, Second Edition (SRS-2) to investigate the phenotypic presentation of social-behavioral impairments for the synaptopathy—*SYNGAP1*-related Intellectual Disability (*SYNGAP1*-ID) (n=32) compared with a phenotypically similar disorder Phelan-McDermid Syndrome (PMD) (n=27) and healthy controls (n=43). A short form SRS-2 analysis (n=85) was also conducted. Both *SYNGAP1*-ID and PMD had significantly elevated total and subcategory T-scores, with no significant score differences between *SYNGAP1*-ID and PMD, consistent between the full and short form. Mild to severe deficiencies in reciprocal social behavior were found in 100% of PMD individuals and 87.1% of *SYNGAP1*-ID individuals. Additionally, the short form demonstrated greater utility for *SYNGAP1-*ID participants due to lower item-omission rates. In conclusion, significant impairment in reciprocal social behaviors is highly prevalent in *SYNGAP1*-ID.

## INTRODUCTION

Genetic mutations affecting the development and function of the neuronal synapse lead to synaptopathies, which frequently include developmental delays or intellectual disability, behavioral dysregulations, epilepsy and social impairments. There are a growing number of genes which encode proteins found at the neuronal synapse that when mutated result in neurodevelopmental or neuropsychiatric disorders. Indeed, large genomic sequencing studies have determined that synaptic genes are one of the more common classes of genes where mutations are a significant risk factor for the development of autism spectrum disorders (Satterstrom et al., 2020). Despite a broad understanding of possible neurologic and psychiatric phenotypes associated with mutations of synaptic genes, detailed phenotypic analyses of these disorders are largely lacking. Here, we investigated the social responsiveness for the synaptopathy – *SYNGAP1*-related Intellectual Disability (*SYNGAP1*-ID) in comparison with another better characterized synaptopathy, Phelan McDermid syndrome (PMD), and healthy controls (HC).

*SYNGAP1*-ID is a genetic disorder caused primarily by de novo loss-of-function single nucleotide variants in *SYNGAP1* or, less commonly, a hemizygous deletion of chromosome 6p21.3, the cytogenic location of *SYNGAP1*(Agarwal et al., 2019; Jimenez-Gomez et al., 2019; Keller et al., 2017; Vlaskamp et al., 2019). This gene encodes for the SynGAP protein, which functions as a GTPase that is essential for synaptic development, structure, function and plasticity (Gamache et al., 2020; Keller et al., 2017). Mutations of *SYNGAP1* increase neurotransmission at excitatory glutamatergic synapses resulting in imbalance of excitatory/inhibitory neurotransmission (Agarwal et al., 2019; Keller et al., 2017; Kozol et al., 2015; Vlaskamp et al., 2019).

PMD is another genetic synaptopathy caused primarily by the deletion of the distal long arm of chromosome 22 (encompassing 22q13), with clinical manifestations remarkably similar to *SYNGAP1*-ID (Phelan, K. & McDermid, H., 2012). The neurological deficits associated with PMD have been linked to haploinsufficiency of *SHANK3*, a gene encoding a protein enriched in the postsynaptic density of excitatory synapses (Phelan, K. & McDermid, H., 2012). Reduction in the SHANK3 protein due to haploinsufficiency of its gene reduces excitatory glutamatergic neurotransmission also resulting in an imbalance of excitatory/inhibitory neurotransmission, although thought to be in the opposite direction due to mutations in *SYNGAP1* (Shcheglovitov et al., 2013; Peca et al., 2011).

Despite mutations in *SYNGAP1* and *SHANK3* resulting in opposite effects on E/I ratio, the phenotypic presentations of both *SYNGAP1*-ID and PMD are similar in that they can include epilepsy, intellectual disability (ID), autism spectrum disorder (ASD), severe expressive and receptive speech delay, hypotonia, sleep abnormalities, global developmental delays, and behavioral issues (Berryer et al., 2013; Jimenez-Gomez et al., 2019; Mignot et al., 2016; Phelan, K. & McDermid, H., 2012; Smith-Hicks et al., 2021; Soorya et al., 2013; Vlaskamp et al., 2019). Autism-related social impairments are commonly reported symptoms in both patient populations. In the largest cohort reported to date, 30/57 (52.6%) of patients with pathogenic *SYNGAP1* mutations were diagnosed with autism spectrum disorder (Vlaskamp et al., 2019). Similarly, in a cohort of 27 participants with *SYNGAP1*-ID, 52% were found to have an ASD diagnosis (Wright et al., 2022). In a study analyzing 32 patients with PMD, 27 (84%) met the criteria for autism spectrum disorder, as assessed by the Autism Diagnostic Interview-Revised and the Autism Diagnostic Observation Schedule-G (Soorya et al., 2013). However, the prevalence and severity of social impairments for children with *SYNGAP1*-ID has yet to be assessed systematically. Moreover, no study to date has compared the severity of social impairment between synaptopathies or with neurotypical children.

Here, we assess the severity of social behavioral impairments resulting from the synaptopathy, *SYNGAP1*-related intellectual disability (*SYNGAP1*-ID) using a validated instrument, the Social Responsiveness Scale Second Edition (SRS-2) (Constantino, J.N. & Gruber, C.P., 2012). We utilized the SRS-2 to quantify the social behavioral phenotypes of participants diagnosed with *SYNGAP1*-ID because it is well-validated in multiple populations and easy and quick to administer. The SRS-2 is a 65-item, Likert-scale, objective measure of autism-associated reciprocal social interactions, typically completed by a caregiver or teacher of a child. It is an extensively cited measure within ASD literature, and is used in practice to screen for ASD, or in clinical settings to detect subtle variations in the severity of symptoms over time, across individuals, or as a treatment outcome measure (Constantino, J.N. & Gruber, C.P., 2012). The raw scores are normalized by gender, age group (preschool, school-age, and adult), and rater identity (parent or teacher). Besides the total severity score, the survey also creates sub-scores for the following categories: social awareness, social cognition, social communication, social motivation, and restricted interests and repetitive behaviors.

Previous studies assessing the psychometric properties of the full SRS-2 in children with severe ID and language impairments suggests that the full SRS-2 is limited in its ability to assess behavioral phenotypes in these individuals (Gergoudis et al., 2020; Sturm et al., 2017). A shortened version of the SRS-2 was created in an effort to limit the influence of age, expressive language, behavior problems and nonverbal IQ on survey scores, while maintaining a unidimensional factor structure (Sturm et al., 2017). This version of the SRS-2 consists of 16 items taken from the full form (Sturm et al., 2017). A study analyzing the psychometric properties of both the full and shortened SRS-2 forms in PMD patients (n=91) found that the shortened SRS-2 showed significant improvement in validity and reliability as compared to the full form for this patient population (Gergoudis et al., 2020). For this reason, we conducted analysis with both the full and shortened versions of the SRS-2 form for participants that completed the School-Age form, also known as the original SRS form.

## METHODS

### Recruitment

Participants for this study were recruited from the Bluebird Circle Clinic for Pediatric Neurology at Texas Children’s Hospital with assistance from the following family advocacy foundations: Phelan McDermid Syndrome Foundation, *SYNGAP1* Foundation and the SynGAP Research Fund, Inc.

Adult caregivers of children younger than 21 years of age with a diagnosis of *SYNGAP1*-ID or PMD were eligible to participate. Participant’s IQ or presence of ASD diagnosis was not ascertained. Exclusionary criteria included individuals greater than 21 years old. Those interested in participating in the study were given information about the study over the phone. Consent forms were reviewed in the same call, and then mailed to the family for signature with a return envelope. Once consent was obtained, the questionnaire was completed over the phone and answers were recorded by a member of the research staff. Questions were read to participants as they were not local. Participants were asked 65 total questions from the relevant SRS-2 questionnaire. For children between the age of 1.5 and 4.5 years old, a Preschool form was completed, and all other subjects (4.5-21 y/o) completed the School Age form. Completing the questionnaire took 15-20 minutes. Caregivers were given the option to leave questions unanswered if not applicable or unknown. However, in accordance with the assessment instructions, if 7 or more items were left blank the survey was deemed unusable.

Once raw scores were collected, the accompanying scoring worksheet was completed by a member of the research staff to obtain treatment subscales and total raw scores. The raw scores were then used to determine T-scores, normalized for age-group. School Age forms also normalized scores for gender and rater identity (which was a parent in all cases). T-scores below 59 were considered within normal limits, and any score above was further subcategorized into the mild (60-65), moderate (66-75), or severe range (≥76). T-scores were obtained for the total score as well as each subscale.

Participating caregivers were also asked to complete the survey for neurotypical siblings to participate as healthy control individuals. The surveys were completed between December 2020 and January 2023.

Ethical approval was obtained from the Institutional Review Board for Baylor College of Medicine and Affiliated Hospitals (H-48239).

### Demographics

Participants ranged from age 2 to 21 years. Table 1 describes the general demographic characteristics of all participants for which data was obtained.

**Table 1.** Demographic characteristics of all subjects

| **Demographic characteristics (n=102)** |  |  |
| --- | --- | --- |
| Age in years (Mean/SD) | 10.1 | 4.98 |
| Sex (n/%) |  |  |
| Male | 51 | 50 |
| Female | 51 | 50 |
| Race/ethnicity (n/%) |  |  |
| Caucasian | 73 | 71.6 |
| Asian | 4 | 3.9 |
| Hispanic | 16 | 15.7 |
| Two or more | 6 | 5.9 |
| Unreported | 3 | 2.9 |

#### Full SRS-2 Analysis

Participants included in the full SRS-2 analysis were those who satisfactorily completed a version of the full SRS form (School Age or Preschool versions). A Kruskal-Wallis test was used to compare medians for age in the three groups. A Chi-squared test was used to determine statistical significance in sex distribution among the three groups. No statistically significant difference was found between age or sex distribution amongst all three groups (Table 2).

**Table 2.** Mean age and gender distributions amongst all three patient groups used for the analysis of the full SRS-2. Kruskal-Wallis statistic=3.07; **Chi-square, df= 2.79, 2

|  | ***SYNGAPI-ID* (n=31)** | **PMD (n=18)** | **HC (n=43)** | **p-Value** |
| --- | --- | --- | --- | --- |
| Age (sd) | 9.0 (5.2) | 11.1 (5.6) | 10.7 (4.3) | 0.22* |
| Female (%) | 58.1 | 33.3 | 48.8 | .25** |
| Male (%) | 41.9 | 66.7 | 51.2 | .25** |

#### Short SRS-2 Analysis

Only subjects who filled out the School Age form were included in the shortened SRS analysis, because the short form was originally derived from the SRS-2 School Age form (Sturm et al., 2017). A total score and item-level analysis was conducted.

No statistically significant difference was found between age or sex distribution amongst all three groups for this analysis group (Table 3). In the short form analysis, participants that failed to answer 2 or more of the 16 items (12.5%) were omitted (n=5). This decision was made based on the SRS-2 manual’s guidelines to omit participants who failed to answer 7/65 (10.7%) of the items on the full form (Constantino, J.N. & Gruber, C.P., 2012).

**Table 3.** Mean age and sex distributions amongst all three patient groups used for analysis of the short SRS survey. *Kruskal-Wallis statistic=0.54; * *Chi-square, df= 0.92, 2

|  | ***SYNGAPI-ID* (n=23)** | **PMD (n=21)** | **HC (n=41)** | **p-Value** |
| --- | --- | --- | --- | --- |
| Age (sd) | 10.3 (4.2) | 11.4 (5.0) | 11.0 (4.1) | 0.76* |
| Female (%) | 60.9 | 42.9 | 48.8 | .63** |
| Male (%) | 39.1 | 57.1 | 51.2 | .63** |

#### Statistical analysis

Raw scores and T scores from surveys were recorded in Microsoft Excel. Descriptive statistics for total scores and subscale scores (social awareness, social cognition, social communication, social motivation, restricted interests and repetitive behavior), both raw and T scores, were performed using GraphPad Prism version 9. Descriptive statistics for item-level responses were also obtained for the short SRS form. Complete analysis of full and short form scores was conducted. A Spearman correlation test was performed to determine the correlation between full and short form scores. For all analyses, to compare the three population groups, the Kruskal-Wallis non-parametric test was used because data did not pass D’Agostino & Pearson normality test. For pairwise comparisons, Dunn’s multiple comparison test was used. All statistical tests were performed using GraphPad Prism version 9.

## RESULTS

### Full Form Analysis

The SRS total scores, both raw and T scores, were significantly increased (p<0.0001) for both *SYNGAP1*-ID and PMD populations as compared to healthy controls (Figure 1 and Table 4). No significant difference was found between the total scores of *SYNGAP1*-ID and PMD patients. Additionally, for all SRS-2 subscale scores (raw and T scores), significant difference (p<0.0001) was found between PMD and *SYNGAP1*-ID participants and healthy control, with no significant difference between PMD and *SYNGAP1*-ID scores. A total of 10 participants were omitted from the full form analysis due to incomplete data sets caused by a number of inapplicable questions for parents. (*SYNGAP1-*ID, n=1; PMD, n=9). Analyses were performed to determine whether total scores were impacted by sex, but results demonstrated no significant difference between total scores based on sex (Supplemental Figure S1).

**Table 4.** Full form raw scores for total and subcategories, mean (SD)

|  | *SYNGAP1*-ID<br>(n=31) | PMD<br>(n=18) | HC<br>(n=43) | Kruskal-Wallis<br>Statistic | KW *p*-<br>Value |
| --- | --- | --- | --- | --- | --- |
| Total Score | 105.1 (29.99) | 116.2 (20.40) | 22.49 (15.26) | 65.03 | <0.0001 |
| Social Awareness | 14.19 (4.12) | 17.50 (2.77) | 3.98 (2.75) | 66.70 | <0.0001 |
| Social Cognition | 19.87 (6.09) | 21.11 (4.91) | 3.86 (2.70) | 65.59 | <0.0001 |
| Social Communication | 35.68 (10.94) | 41.28 (7.70) | 6.37 (5.42) | 66.66 | <0.0001 |
| Social Motivation | 12.65 (5.65) | 13.06 (5.59) | 5.19 (4.32) | 34.40 | <0.0001 |
| Restricted Interests & Repetitive Behavior | 22.71 (8.01) | 23.44 (6.19) | 3.00 (2.98) | 63.92 | <0.0001 |
| DSM-5 SCI | 82.39 (23.53) | 92.94 (16.16) | 19.49 (13.06) | 64.32 | <0.0001 |

**Fig. 1.**
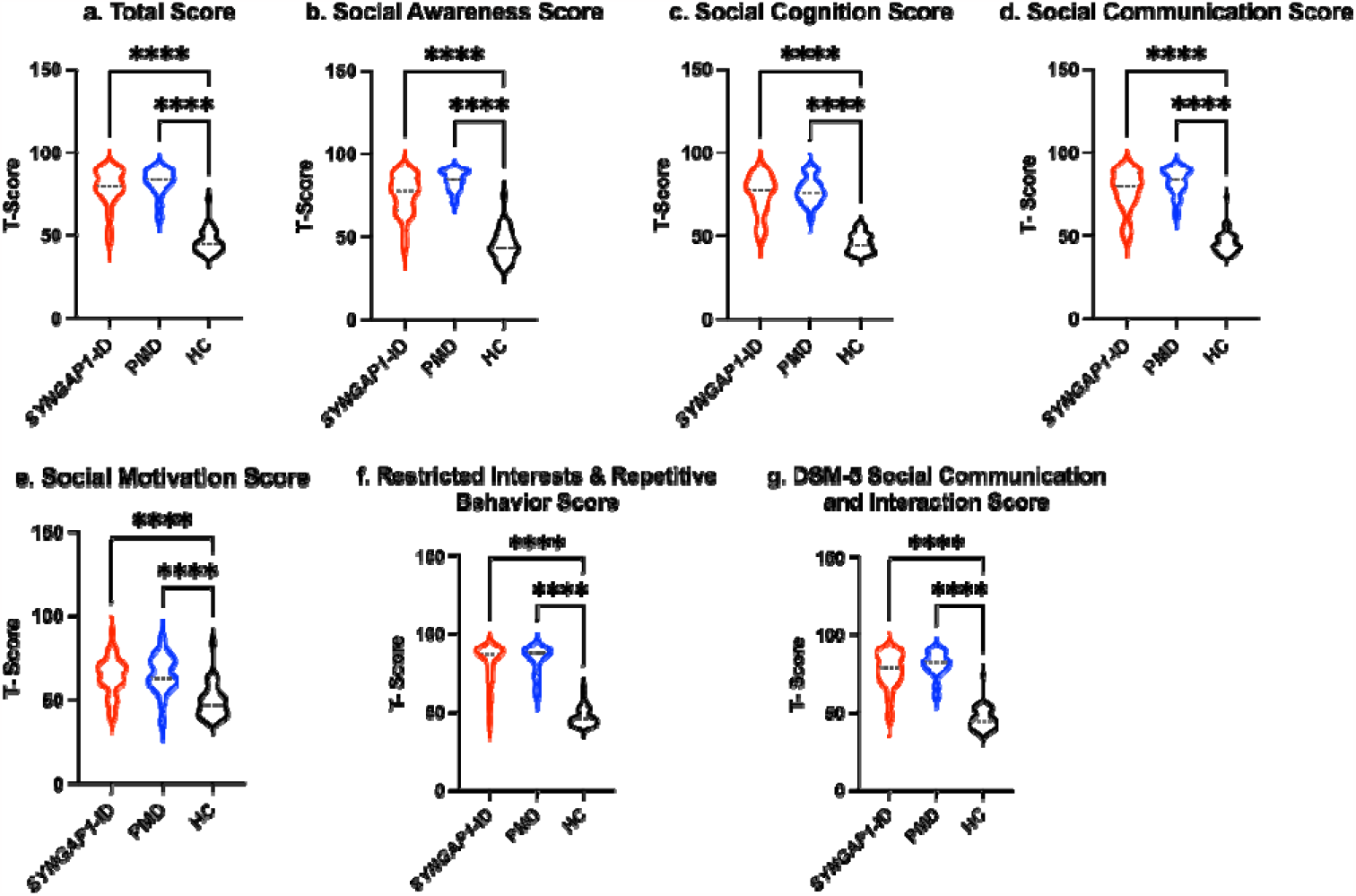
Total T-scores (a) and subscale T-scores for *SYNGAP1*-related intellectual disability (*SYNGAP1*-ID), Phelan-McDermid Syndrome (PMD), and healthy controls (HC). Subscales: (b) Social Awareness, (c) Social Cognition, (d) Social Communication, (e) Social Motivation, (f) Restricted Interests and Repetitive Behavior, (g) DSM-5 Social Communication and Interaction Score. Dunn’s multiple comparison test **** p<0.0001. Thick dash lines represent medians and thin dashed lines represent quartiles

Based on the SRS full form cut-offs, 100% of PMD individuals and 87.1% of *SYNGAP1*-ID individuals met the criteria for mild to severe deficiencies in reciprocal social behavior, as defined by the SRS scale (Table 5). The most common category was “Severe” deficiencies with 83.3% of PMD individuals and 67.7 % of *SYNGAP1*-ID individuals falling into this category. A small percentage of *SYNGAP1*-ID individuals (12.9%) fell in the “Within normal limits” category. These data suggest that impairments in reciprocal social behavior is very common for individuals with these synaptopathies, with a majority presenting with severe impairments.

**Table 5.** Labels as defined by SRS Scale (n/%); Within normal limits (59T or below), Mild range (60T-65T), Moderate range (66T-75T), Severe range (76T or higher)

|  | *SYNGAP1*-ID<br>(n=31) | PMD<br>(n=18) | HC<br>(n=43) |
| --- | --- | --- | --- |
| Within Normal Limits | 4 (12.9) | 0 (0) | 42 (97.7) |
| Mild | 1 (3.2) | 1 (5.5) | 0 (0) |
| Moderate | 5 (16.1) | 2 (11.1) | 1 (2.3) |
| Severe | 21 (67.7) | 15 (83.3) | 0 (0) |

### Short form analysis

An analysis of participants who satisfactorily completed a School Age form (n=85) showed that both *SYNGAP1*-ID (n=23) and PMD (n=21) individuals had significantly elevated (p<0.0001) total scores compared to healthy controls (n=41) (Figure 2). A total of 5 participants were omitted from the total score short form analysis due to missing data. Additionally, both *SYNGAP1*-ID and PMD individuals had significantly elevated (p<0.0001) scores as compared to healthy controls in every item, except item 42 (Table 6). No significant differences were found between scores of *SYNGAP1*-ID and PMD individuals in any of the items or in total score. Average scores from all items on the short form were obtained and are displayed in Table 6. Item 42 scores showed a statistically significant difference between *SYNGAP1*-ID and HC, but a nonsignificant difference between PMD and HC. Item 42 stated: “Seems overly sensitive to sounds, textures, or smells.” T-scores could not be calculated for the short form, so all analyses were performed with raw scores.

**Table 6.**
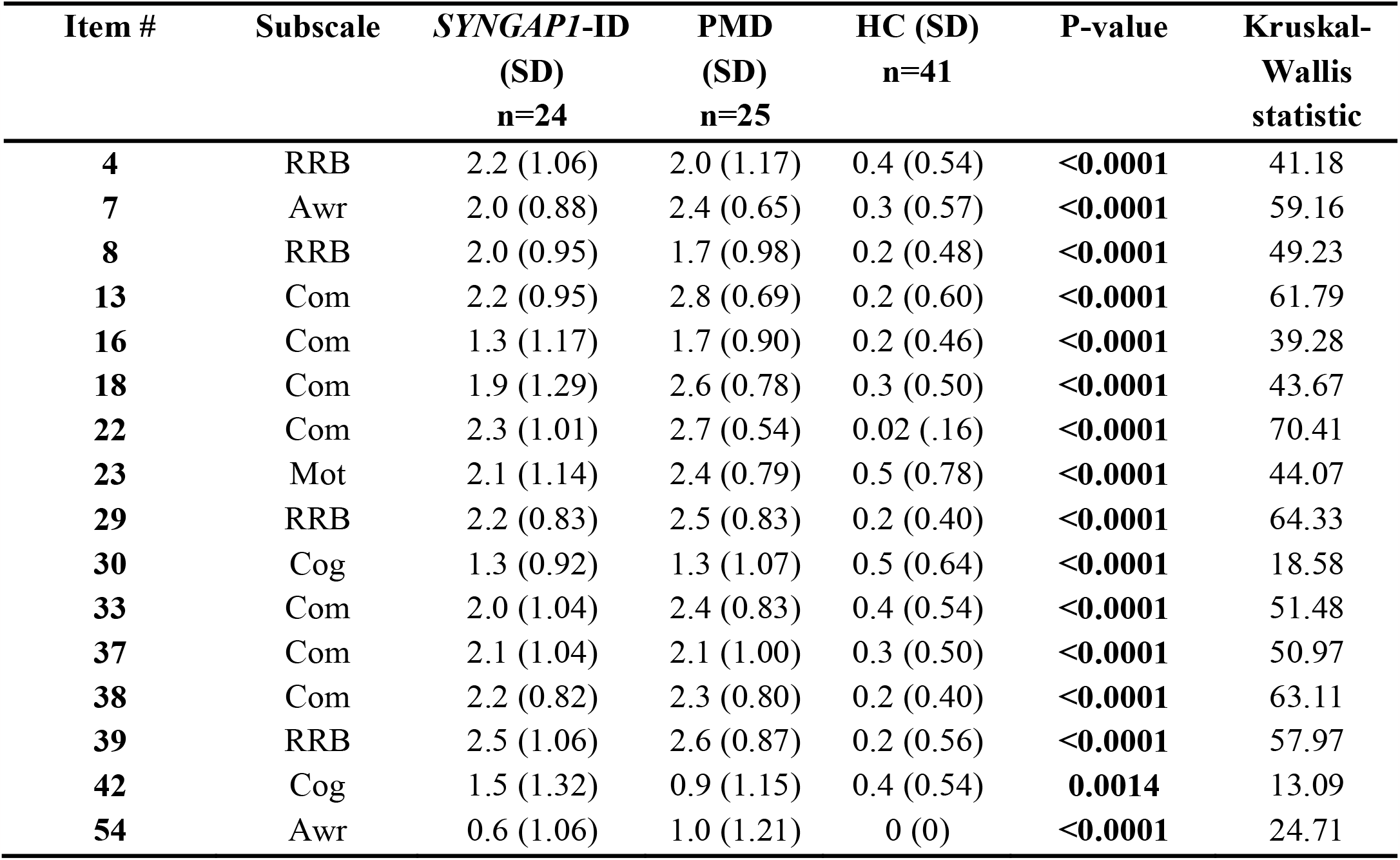
Item-level scores for short form, mean (SD). The number of responses varied for the following items: 13 (PMD, n=22; *SYNGAP1*-ID, n=23), 18 (PMD, n=18; *SYNGAP1*-ID, n=23), 23 (PMD, n=23; *SYNGAP1*-ID, n=23), 29 (PMD, n=24), 33 (PMD, n=24; *SYNGAP1*-ID, n=23). Abbreviations: Awr = Social Awareness; Cog = Social Cognition; Com = Social Communication; Mot = Social Motivation; RRB = Restricted Interests and Repetitive Behavior

| Item # | Subscale | *SYNGAPI*-ID<br>(SD)<br>n=24 | PMD<br>(SD)<br>n=25 | HC (SD)<br>n=41 | P-value | Kruskal-Wallis<br>statistic |
| --- | --- | --- | --- | --- | --- | --- |
| 4 | RRB | 2.2 (1.06) | 2.0 (1.17) | 0.4 (0.54) | <0.0001 | 41.18 |
| 7 | Awr | 2.0 (0.88) | 2.4 (0.65) | 0.3 (0.57) | <0.0001 | 59.16 |
| 8 | RRB | 2.0 (0.95) | 1.7 (0.98) | 0.2 (0.48) | <0.0001 | 49.23 |
| 13 | Com | 2.2 (0.95) | 2.8 (0.69) | 0.2 (0.60) | <0.0001 | 61.79 |
| 16 | Com | 1.3 (1.17) | 1.7 (0.90) | 0.2 (0.46) | <0.0001 | 39.28 |
| 18 | Com | 1.9 (1.29) | 2.6 (0.78) | 0.3 (0.50) | <0.0001 | 43.67 |
| 22 | Com | 2.3 (1.01) | 2.7 (0.54) | 0.02 (.16) | <0.0001 | 70.41 |
| 23 | Mot | 2.1 (1.14) | 2.4 (0.79) | 0.5 (0.78) | <0.0001 | 44.07 |
| 29 | RRB | 2.2 (0.83) | 2.5 (0.83) | 0.2 (0.40) | <0.0001 | 64.33 |
| 30 | Cog | 1.3 (0.92) | 1.3 (1.07) | 0.5 (0.64) | <0.0001 | 18.58 |
| 33 | Com | 2.0 (1.04) | 2.4 (0.83) | 0.4 (0.54) | <0.0001 | 51.48 |
| 37 | Com | 2.1 (1.04) | 2.1 (1.00) | 0.3 (0.50) | <0.0001 | 50.97 |
| 38 | Com | 2.2 (0.82) | 2.3 (0.80) | 0.2 (0.40) | <0.0001 | 63.11 |
| 39 | RRB | 2.5 (1.06) | 2.6 (0.87) | 0.2 (0.56) | <0.0001 | 57.97 |
| 42 | Cog | 1.5 (1.32) | 0.9 (1.15) | 0.4 (0.54) | 0.0014 | 13.09 |
| 54 | Awr | 0.6 (1.06) | 1.0 (1.21) | 0 (0) | <0.0001 | 24.71 |

**Fig. 2.**
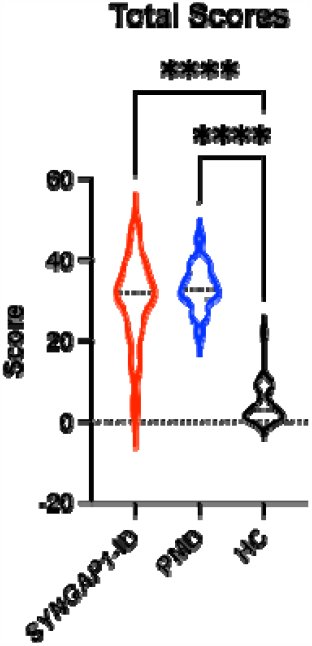
Short SRS form total raw scores for *SYNGAP1*-related intellectual disability (*SYNGAP1*-ID), Phelan-McDermid Syndrome (PMD), and healthy controls (HC). Dunn’s multiple comparison tests **** p<0.0001. Thick dash lines represent medians and thin dashed lines represent quartiles

A Spearman correlation test demonstrated a strong correlation of full form total scores to short form total scores (n=0.9345, Supplemental Figure S2).

## DISCUSSION

Social behavioral abnormalities are commonly reported in individuals with *SYNGAP1*-ID; however, they have not previously been investigated systematically. Here, we administered the validated Social Responsiveness Scale-2 (SRS-2) to determine whether this assessment could appropriately characterize social behavioral phenotypes of individuals with this disorder. Individuals with *SYNGAP1*-ID have elevated total and sub-section scores as compared to healthy controls and indistinguishable impairment compared with those diagnosed a similar monogenic synaptopathy, PMD. Moreover, the vast majority (>85%) of individuals with *SYNGAP1*-ID had a total score greater than the standard cut-off for abnormal social interactions with more than the majority falling into the severe category. Thus, we demonstrate for the first time that reciprocal social interactions are abnormal based upon a standardized measurement.

While the results of the survey demonstrated scores with detectable differences from healthy controls, some participants were omitted from analysis due to incomplete surveys (10.9%). A significant portion (90%) of individuals omitted were individuals with PMD.

The shortened form was created by Sturm et al. to better assess individuals with severe intellectual disability. The three most commonly (n>15) omitted items from our populations (10,15, 62) on the full form were all removed in the short form, demonstrating its utility in these populations. However, a portion of individuals (5.75%) were still omitted from the short form analysis because they failed to answer 2 or more of the 16 items. Commonly omitted items from the shortened form were items 18 and 13 (n=8,5). Table 7 overviews the item content of commonly omitted survey items.

**Table 7.** Commonly omitted items on the School-Age form

| Item # | Item Content | Omissions |
| --- | --- | --- |
| 10 | Takes things too literally and doesn't get the real meaning of a conversation. | 16 |
| 13 | Is awkward in turn-taking interactions with peers (for example, doesn't seem to understand the give-and-take of conversations). | 5 |
| 18 | Has difficulty making friends, even when trying his or her best. | 8 |
| 51 | Has difficulty answering questions directly and ends up talking around the subject. | 19 |
| 62 | Gives unusual or illogical reasons for doing things. | 18 |

No previous study has recorded the use of the SRS survey in *SYNGAP1*-ID patients, however one previous study has assessed the psychometric properties of the full and short SRS survey in PMD patients. Gergoudis et al. found that the full SRS-2 has limited efficacy in the PMD population, and that the shortened form is a more reliable and valid alternative (Gergoudis et al., 2020). This previous study found that many items on the full form have limited relevance and utility for the PMD population, which is resolved in the shortened form. Our data similarly supports the use of the shortened SRS form as compared to the full form in this patient population. Additionally, the strong correlation between the full form and short form total scores adds confidence for the use of the shortened form. However, the omission of 4/25 (16%) of PMD individuals as compared to 1/24 (4.2%) of *SYNGAP1*-ID individuals suggests that even the short form has limitations for the PMD population.

Although no significant differences were found between the scores of PMD and *SYNGAP1*-ID individuals in either the long or short SRS form, the high rate of item-omission in the PMD population differs drastically from the *SYNGAP1*-ID population. Reasons for this observation may relate to the level of non-social behavioral symptom severity in PMD populations as compared to those with *SYNGAP1*-ID, making it difficult for caregivers of these individuals to provide answers to specific items. The omission-rate discrepancies between PMD and *SYNGAP1*-ID individuals warrants further investigation.

This study was limited by a relatively small sample size that resulted in using non-parametric statistical tests to analyze data, yielding less power than their parametric counterparts.

## CONCLUSION

By utilizing the Social Responsiveness Scale assessment, we discovered the majority of individuals with *SYNGAP1*-related intellectual disability (*SYNGAP1*-ID) have severe impairments in reciprocal social behavior similar to individuals with a related synaptopathy-Phelan McDermid Syndrome (PMD). We also found that the shortened SRS survey by Sturm et al. is a more feasible assessment for use in children with these synaptopathies because of lesser influence of confounding factors, such as NVIQ, on scores. However, differences in omission rate between *SYNGAP1*-ID and PMD individuals point to lower utility of the assessment for individuals with PMD. We propose that the shortened SRS form can be utilized to characterize social behavioral phenotypes in patients with *SYNGAP1*-ID and has less utility for those with PMD.

## Data Availability

All data produced in the present study are available upon reasonable request to the authors

## SUPPLEMENTAL DATA

**Figure S1.**
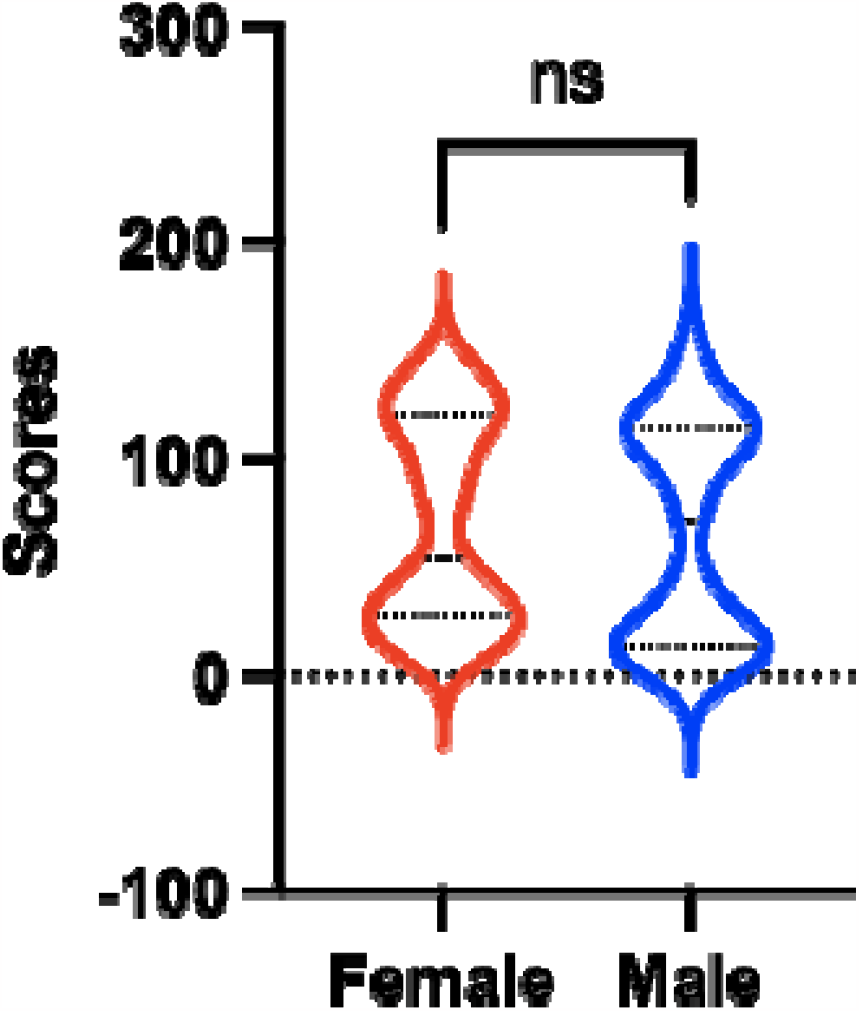
Total scores on full form for males (n=47) and females (n=45). Significance tested using a Mann-Whitney test (p = 0.2324)

**Figure S2.**
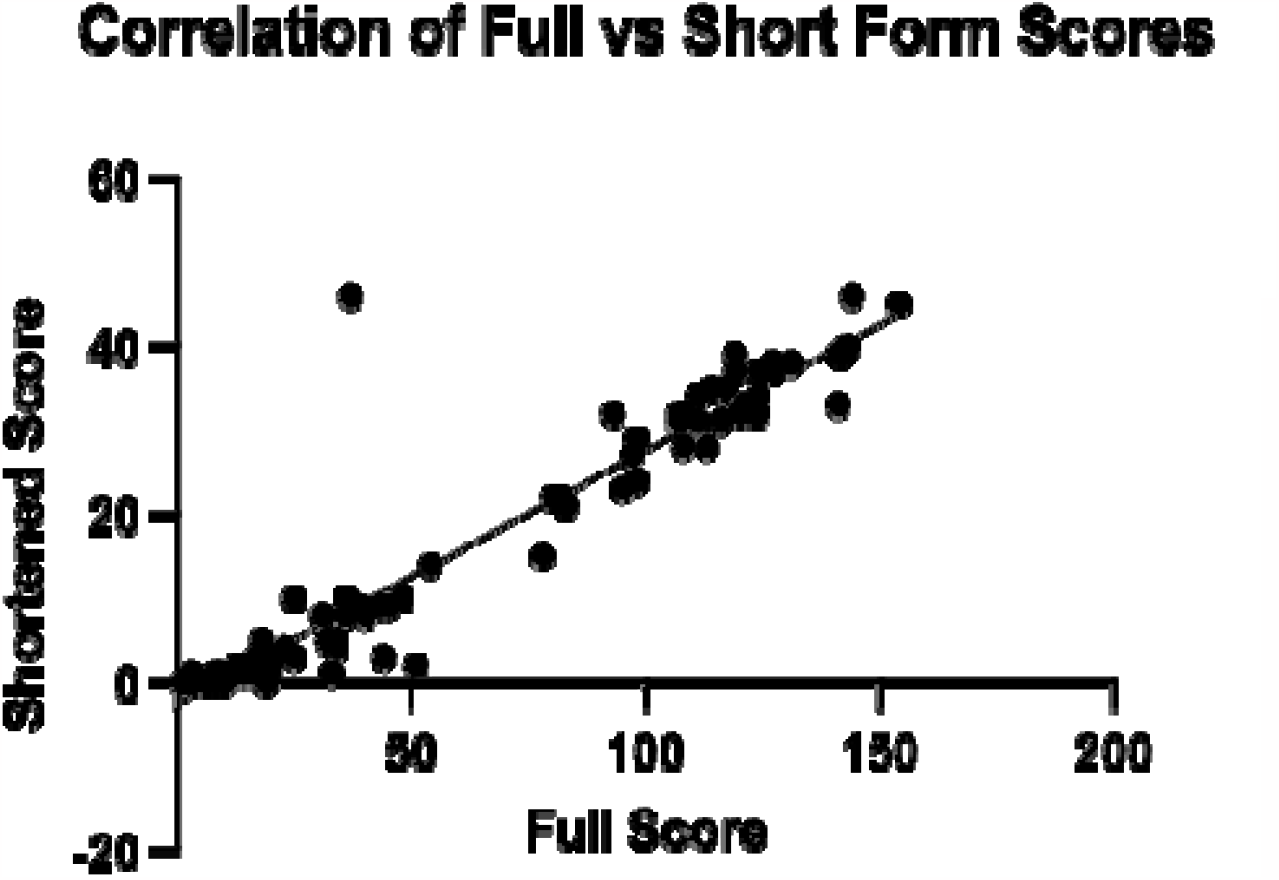
Simple linear regression of full form total scores vs short form total scores (n=81); R^2 = 0.8911

